# Machine learning assisted discovery of synergistic interactions between environmental pesticides, phthalates, phenols, and trace elements in child neurodevelopment

**DOI:** 10.1101/2023.02.02.23285222

**Authors:** Vishal Midya, Cecilia Sara Alcala, Elza Rechtman, Irva Hertz-Picciotto, Chris Gennings, Maria Rosa, Damaskini Valvi

## Abstract

A growing body of literature suggests that higher developmental exposure to individual or mixtures of environmental chemicals (ECs) is associated with autism spectrum disorder (ASD). However, the effect of interactions among these ECs is challenging to study. We introduced a composition of the classical exposure-mixture Weighted Quantile Sum (WQS) regression, and a machine-learning method called signed iterative random forest (SiRF) to discover synergistic interactions between ECs that are (1) associated with higher odds of ASD diagnosis, (2) mimic toxicological interactions, and (3) are present only in a subset of the sample whose chemical concentrations are higher than certain thresholds. In the case-control Childhood Autism Risks from Genetics and Environment study, we evaluated multi-ordered synergistic interactions among 62 ECs measured in the urine samples of 479 children in association with increased odds for ASD diagnosis (yes vs. no). WQS-SiRF discovered two synergistic two-ordered interactions between (1) trace-element cadmium(Cd) and alkyl-phosphate pesticide - diethyl-phosphate(DEP); and (2) 2,4,6-trichlorophenol(TCP-246) and DEP metabolites. Both interactions were suggestively associated with increased odds of ASD diagnosis in a subset of children with urinary concentrations of Cd, DEP, and TCP-246 above the 75^th^ percentile. This study demonstrates a novel method that combines the inferential power of WQS and the predictive accuracy of machine-learning algorithms to discover interpretable EC interactions associated with ASD.

**Synopsis:** The effect of interactions among environmental chemicals on autism spectrum disorder (ASD) diagnosis is challenging to study. We used a combination of Weighted Quantile Sum regression and machine-learning tools to study multi-ordered synergistic interactions between environmental chemicals associated with higher odds of ASD diagnosis.

**Graphical Abstract:** 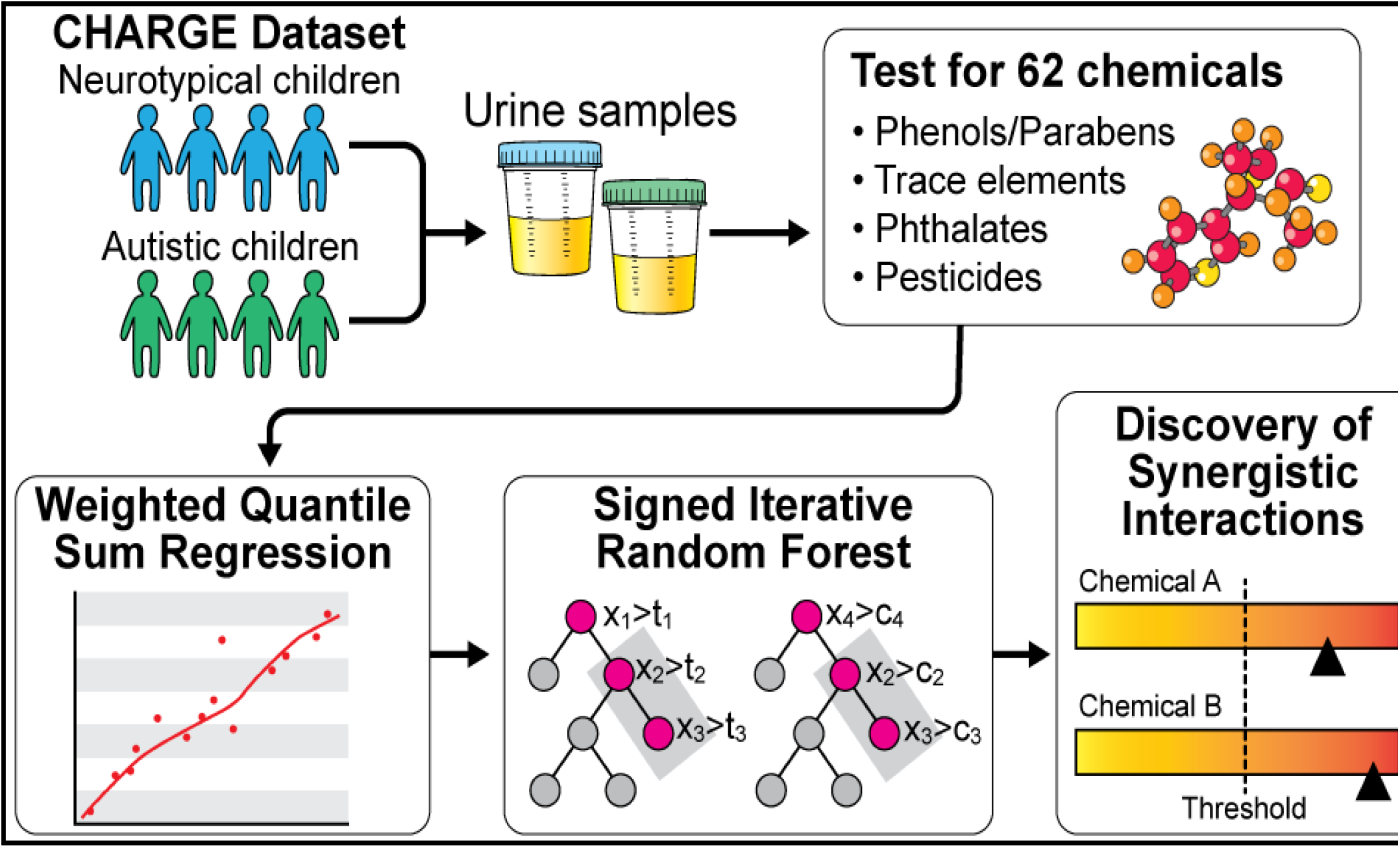

## Introduction

Autism spectrum disorder (ASD) is a neurodevelopmental disorder characterized by deficits in social communication and interaction and repetitive and stereotyped interests and behaviors^1^. ASD prevalence has increased drastically recently and is a public health concern worldwide. According to the Centers for Disease Control program Autism and Developmental Disabilities Monitoring (ADDM) Network, approximately 1 in 44 children have been diagnosed with ASD^2, 3^. In the past decade, a growing number of epidemiological studies have associated early-life environmental exposures with ASD^4^. These environmental exposures include air pollution^5-9^, nutrition, and environmental chemicals like volatile organic compounds, solvents, and endocrine-disrupting chemicals (EDCs). Among multiple EDCs, the field of metals exposure and ASD research is continuously evolving^10, 11^, suggesting a compelling link between higher inorganic metalloid arsenic and ASD in children^12^. The association between trace elements and other endocrine-disrupting chemicals, such as BPA, and parabens, with ASD has been understudied^13^; however, studies have shown that they are potential risk factors for child behavioral outcomes^14-16^.

Although the exact cause of ASD remains unclear, research on its etiology has suggested an interplay of multiple genetic and early environmental contributions that differ between individuals^4, 17, 18^. Genetic and environmental factors may impact typical brain development, including neuron formation and migration, synapse formation, or neural connectivity, ultimately leading to ASD^4^. Environmental chemical exposures biologically interact through pathophysiologies, including the direct disruption of cells and structures of the nervous system, endocrine hormone- or immune system-mediated impacts, and epigenetic changes, to name a few^4^. However, there is a lack of environmental epidemiology studies assessing potential chemical-chemical interactions in ASD. Among the very few studies, Curtin et al. examined whether the dynamic interaction of zinc-copper cycles, which regulate metal metabolism, are disrupted in ASD^19^. Findings showed that the interaction between cyclical co-occurrence between zinc and copper is disrupted in ASD^19, 20^.

The concept of “interaction” has been construed in many ways through different scientific fields^21^. For example, in current epidemiological studies, interactions are usually reported through association estimates of their effect sizes or inclusion probabilities^22-33^. Though estimating associations is essential, most methods do not provide any mechanistic or biological insight, possibly because the reported interactions are of particular functional forms (for example, multiplication of exposures) rather than representing their collective activities beyond certain concentration thresholds^34^. Further, after applying certain dimension reductions, most interactions are reported between sets of reduced exposures, limiting interpretability. In addition, such interactions provide a population-level estimate, with each sample providing some contribution to the overall estimate.

On the other hand, the toxicological representation of interactions is easier to comprehend. Through the collective activities of the chemicals, (1) one can identify the mechanism of synergistic or antagonistic behavior that might arise beyond the concentration thresholds (and not just the regression coefficient of multiplicative associations), and (2) the use of concentration thresholds reflect the toxicological underpinning of classical threshold based chemical dose-response studies^35-37^. Moreover, as the number of chemical exposures increases, searching for multi-ordered interactions gets computationally very intensive. Most current methods, therefore, “hard code” or pre-specify interaction terms in models, but such strategies are limited due to restrictions on sample size and are usually underpowered^38, 39^. In comparison, Kernel Machine Regression or Bayesian factorization-inspired methods discover interactions with certain functional forms that do not represent any collective activity or concentration thresholds^30, 32, 40^. The lack of similarity with toxicological threshold-based dose-response studies makes it difficult to find any biologically relevant interpretation of the recovered interactions. Note that such interactions can only be present in a subset of the population since not every sample will have chemical concentrations beyond certain thresholds.

As a possible alternative to address this problem of interpretability, tree-based machine learning (ML) models provide a natural solution to represent collective activities of chemical exposures as threshold-based interactions. Nevertheless, a significant challenge was that most of these tree-based models were black-box, creating tension between prediction quality and meaningful biological insight. Moreover, a predictive machine-learning model might not be the optimal model for inference ^41^. However, in recent epidemiological studies, interpretable tree-based machine-learning tools were used to discover simultaneously co-occurring chemicals, similar to classical Weighted Quantile Sum (WQS) Regression models ^42-46^. Separately in computational biology, using a novel ML algorithm called random intersection trees^47^, Basu et al.^34, 48^ introduced the “signed iterative random forest” (SiRF) algorithm to discover interactions through collective activities. Moreover, SiRF can efficiently search for the few stable and highly occurring interactions instead of going through each possible interaction term. Since exposure to environmental chemicals occurs simultaneously, we intend to use a combination of the WQS regression and the ML method Signed Iterative Random Forest to search for interactions that mimic toxicological interactions. Using data from the Childhood Autism Risks from Genetics and Environment (CHARGE) study, we aimed to identify multi-ordered synergistic interactions between environmental chemicals at specific exposure thresholds associated with higher odds of ASD. We further examined whether the directionality of the interactions remained unaltered even after adjusting for the potential effects of the overall chemical mixtures.

## Methods

### Study Design and Population

Details about the CHARGE study have been reported in Bennett et al.^13^ Briefly, the Childhood Autism Risks from Genetics and Environment (CHARGE) is a case-control study that recruited between 2006 and 2017 three groups of children: (1) children with ASD (2) children with developmental delay (DD) but not ASD, and (3) children with typical development (TD)^49^. Children from the first two groups were mainly identified from the California Department of Developmental Services. The department coordinates services for individuals with developmental disabilities and is inclusive of all residents of California regardless of their place of birth, religion, or financial resources ^13^. The third group (controls) was sampled from California birth files utilizing frequency matching of ASD cases comprised of the following characteristics, age, sex, and broad geographic regions up to 10 counties. Children from all three groups were: a) aged 24-60 months at recruitment; b) living with a biological parent who speaks English or Spanish; c) born in California; and d) residing in the study catchment area. CHARGE study included all children with at least 16 mL of urine collected at their assessment and available for chemical analysis. In addition, detailed demographic characteristics of the parents and children were collected during the study visit. However, in this present study, we only included children with either ASD (from group 1) or typical development (from group 3), totaling a sample size of 479.

### Exposure Assessment

We collected a single urine sample from each participating child during their visit. All samples were frozen immediately at -20□C and remained frozen until analysis. The samples were shipped on dry ice to the New York State Department of Health’s (NYSDOH) Wadsworth Center’s Human Health Exposure Analysis Resource (HHEAR) Targeted Analysis Laboratory for analysis. Enzymatic deconjugation and liquid-liquid extraction were used to assess the specific phenolic compounds previously described ^50, 51^. A comprehensive description of the exposure assessment of the targeted phenolic compounds can be found in Bennett et al.^13, 50-52^. Urinary phthalate metabolites (PhMs) were analyzed using enzymatic deconjugation, solid-phase extraction (SPE), and an isotope dilution method of quantification^53^. Further information on the analysis of the PhMs is explained elsewhere^13, 53, 54^. We used the SPE method and the HPLC-MS/MS to analyze the urine samples for six dialkyl phosphate metabolites (DAPs) described in Bennett et al.^13, 55^. Trace elements were analyzed from urine specimens using the biomonitoring methods based on the ICP-MS at the Laboratory of Inorganic and Nuclear Chemistry at the Wadsworth Center^13, 56^.

Using the following formula, we corrected for specific gravity (SG) urinary concentrations P_c_= Px[(SG_p_-1)/(SG-1)]^57^. P_c_ was the SG corrected metabolite concentration (ng/mL), and SG was the specific gravity of the urine sample. The median specific gravity of the CHARGE participants was 1.0223 ng/mL (SG_p_). In the event that the specific gravity correction factors were greater than 2, they were assigned a value of 2. For values below 0.5, they were assigned 0.5^13^.

### Developmental Assessment

During the study visit, an assessment of ASD diagnosis was conducted (to confirm the diagnosis of ASD indicated during the CHARGE enrollment process) using two gold standard psychometric instruments: the Autism Diagnostic Interview-Revised (ADI-R)^58-60^ and the Autism Diagnostic Observation Schedules (ADOS)^61^. The ADI-R is a semi-structured interview administered to the primary caregiver to diagnose autism and to differentiate autism from other developmental disorders^60^. The ADOS is a semi-structured, standardized assessment where the researcher observes the social interaction, communication, play, and imaginative use of materials by children suspected of having ASD^13, 61^. We utilized the DSM-5 and followed standardized procedures from the ADOS and ADI-R to assign the final diagnosis of ASD^62^. Children from all three groups were administered the Mullen Scales of Early Learning (MSEL) and the Vineland Adaptive Behaviors Scores (VABS)^13^. To confirm that a child did not have ASD, we used the Social Communications Questionnaire to screen for ASD in children in both the developmental delay and general population groups^63^. If a child was positive, we administered the ADI-R and ADOS to determine if they had ASD. All other children enrolled because of a community diagnosis of ASD or DD, but were not confirmed for either of these two diagnoses, were grouped together as Other Early Concerns (OEC)^13^. Children were classified as TD and enrolled as general population controls who did not meet the criteria for either ASD or DD. All classification groups are mutually exclusive. All clinicians participating in the study spoke English and/ or Spanish. Additionally, they achieved research reliability on all of the instruments they administered^13^.

### Statistical Analysis

We used the Weighted quantile sum (WQS)^26^ regression to model the adverse mixture effect of chemicals while simultaneously (1) accommodating the correlation structure of the chemicals and (2) controlling for covariates. Similar to Bennet et al., we conducted this analysis by focusing on the positive association (i.e., adverse directionality) between chemical exposures and ASD status.

To reduce spurious co-occurrences of chemicals, interactions were searched on top of the chemical-mixture effect. A conceptual schematic of different kinds of interactions was presented in Figure 1. Briefly, these interactions mimic classical toxicological interactions where interaction occurs only if the concentration of certain chemicals is above some thresholds. Conceptually, a usual multiplicative interaction between two chemicals (say, A and B) can be mapped to four toxicological interactions, (1) the concentration of A is high, and the concentration of B is high, (2) the concentration of A high, and the concentration of B is low, (3) the concentration of A is low, and the concentration of B is high, and (4) the concentration of A is low, and the concentration of B is low (see Figure 1A). Note that each of the four components is easier to interpret and could directly imply plausible biological interpretation. Moreover, a positive association with multiplicative interaction does not necessarily imply synergy since the higher value of multiplicative interaction does not imply that the concentrations of individual chemicals are also high. However, such a problem of interpretability does not arise for toxicologically mimicked interactions (Figures 1B and 1C). Lastly, multiplicative interactions provide a population-level interaction estimate – where all individuals contribute, whereas the mimicked toxicological interactions are only present in a subset of the population. In the following analysis, we searched for synergistic interactions in the adverse direction, i.e., chemical exposures higher than certain concentration thresholds, mimicking a toxicological interaction.

**Figure 1:**
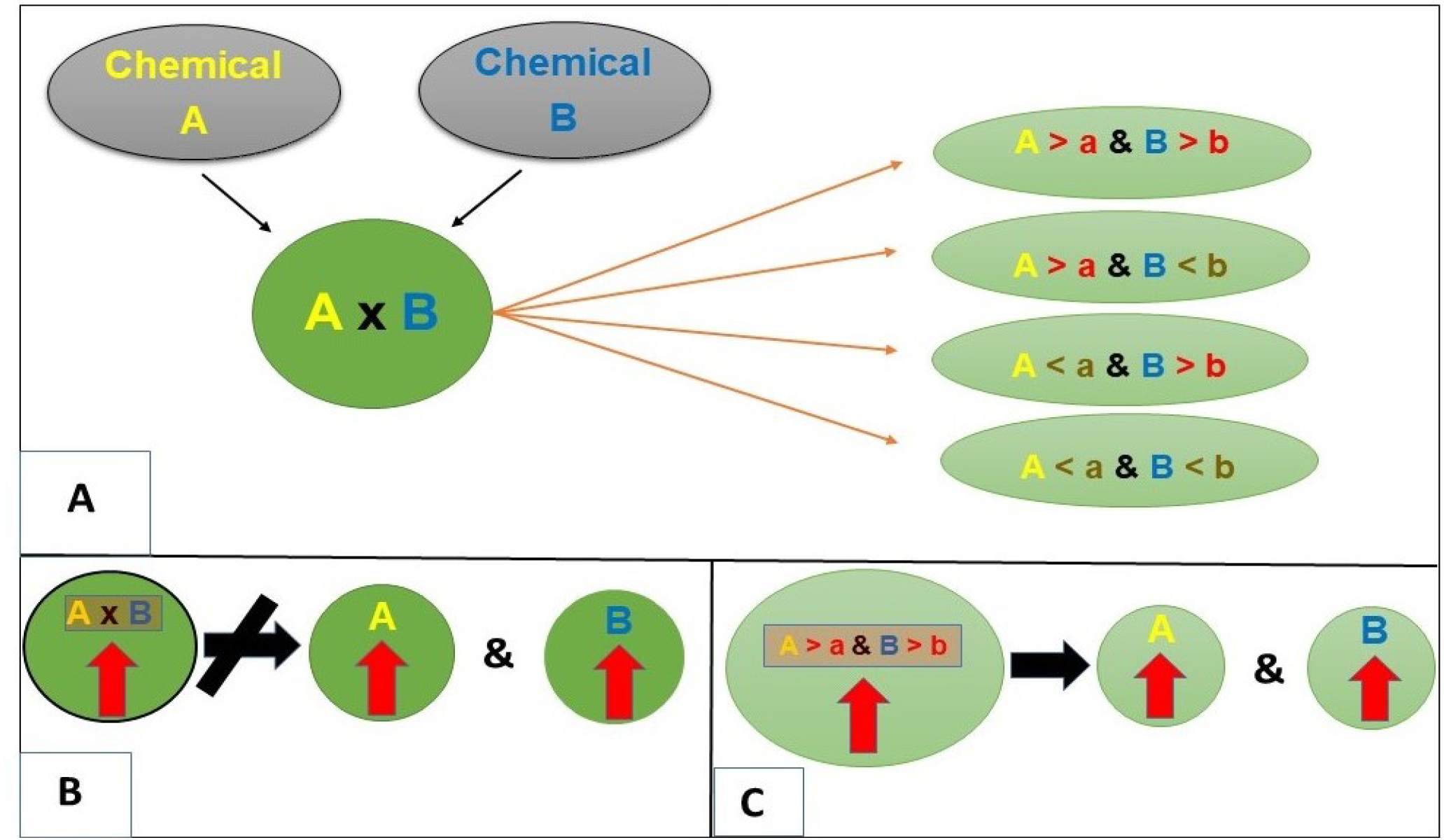
Conceptual illustration of multiplicative and toxicologically mimicked interactions.

The model was controlled for the child’s sex, year of birth, race/ethnicity, age at enrollment, maternal age at the time of childbirth, maternal metabolic conditions during pregnancy (any hypertensive disorder, including obesity or any diabetes), and parental homeowner status (as a proxy of socioeconomic status). These covariates were chosen apriori based on the previous analysis in Bennet et al. ^13^ To make the analysis robust, we implemented the random subset and repeated holdout ^64, 65^ variants of WQS. Assuming the main chemical-mixture effect and the synergistic interactions are additive, we extracted the Pearson residuals from this model and treated the residual as the new outcome (the Pearson residual possesses asymptotic normality)^37, 66^. Therefore, we searched for synergistic interactions on the residuals after adjusting the first-order main mixture effect and the covariates.

We searched for interactions through signed-iterated Random Forest (SiRF), where the Pearson residuals from WQS were the outcome, and the 62 chemicals were the exposures. The SiRF utilizes a combination of state-of-the-art machine-learning tools, iterative Random Forests (iRFs), and recently developed Random Intersection Trees (RITs) to search for interactions within a certain proportion of samples^34, 47, 48, 67^. Instead of searching through all possible combinations, SiRF searches for combinations of exposures prevalent on the decision paths of the generated iRFs. Briefly, we explain how SiRF searches for high-order chemical exposure interactions. First, the model begins with fitting the RF model and reweighting the important exposures. Using the reweighted exposures, multiple RF models are fitted iteratively to reduce the dimensionality of the exposure space without removing marginally unimportant exposures. Second, decision rules are extracted from the iterated RF and fed to a generalization of the RIT to efficiently discover high-order interactions from the decision paths. Last, a bagging step is introduced in the algorithm to assess the “stability” of the recovered interactions through a large number of bootstrapped iterations. Here stability implies the number of times an interaction is detected throughout the iterations; therefore, the higher the recovery rate, the better. Since SiRF searches through particular decision branches, it can incorporate meaningful directionality (in the current study, synergism) while recovering the interactions. The combination of WQS-SiRF can robustly search for interactions without the need to depend on p-values.

In the SiRF part, the model was trained on a subset of data, and then bagging was introduced on the remaining held-out testing data. Therefore, to obtain robust results against the sensitivity of data partitioning, we chose three different data partitions, (1) 70% for training and 30% for testing, (2) 75% for training and 25% for testing, and (1) 80% for training and 20% for testing. Finally, we chose only those interactions with (1) more than 50% stability score and were (2) common to all three data partitioning results. Since the discovered interactions were based on thresholds, they were only present in certain portions of the samples. However, SiRF does not directly estimate the thresholds by itself. Therefore, we created interaction indicators based on their joint concentrations to denote the presence or absence of interactions. For example, if the specific gravity-adjusted concentrations of the chemicals were more than the 75^th^ percentile, then the interaction indicator would be non-zero; else, it would be zero. We created another set of indicators based on the 67^th^ percentile threshold for sensitivity analysis. For WQS analysis, (1) we converted all chemical exposures to deciles, and (2) we included all chemicals irrespective of their percentage detected above LOD. Note that the conversion in deciles for chemical exposures and the growing many decision trees through bootstraps protect against outlying and influential observations.

As sensitivity analyses, (1) we repeated the WQS-SiRF algorithm with data partitioned in 75% for training and 25% for testing without chemicals whose % of detection above LOD was less than 60%, (2) we gradually increased the number of bootstraps, from 250, 500, to 1000, (3) we used the whole dataset to test the model trained on the 75% data, and (4) repeated SiRF to obtain interactions observed in the primary analysis after randomly permuting the ASD status. For descriptive analysis, we calculated the Pearson correlation matrices of log-transformed and specific gravity-corrected 62 chemicals exposures for ASD and TD children. Missing data in covariates was minimal (< 5%) and were imputed using the R package “mice” ^68^. A two-tailed p-value less than alpha at 0.05 is considered statistically significant. All data were analyzed in R version 4.1.2. The detailed mathematical exposition of the algorithm can be found in^48^. In addition, the tuning parameters in WQS-SiRF and random seeds for training and testing data are provided in the supplemental materials.

## Results

The list of all 62 chemicals was presented in Supplemental Table S1, and their LODs (and % detected above LOD) were presented in Supplemental Table S2. Supplemental Table S3 presents the log-transformed and specific-gravity-corrected urinary concentrations of all 62 chemicals for ASD and TD children. Among 62 chemicals, 42 had more than 60% detection rate above LOD (Supplemental Table S2). The specific gravity-adjusted concentration levels and the correlation matrices of the chemicals were presented in Figure 2.

**Figure 2:**
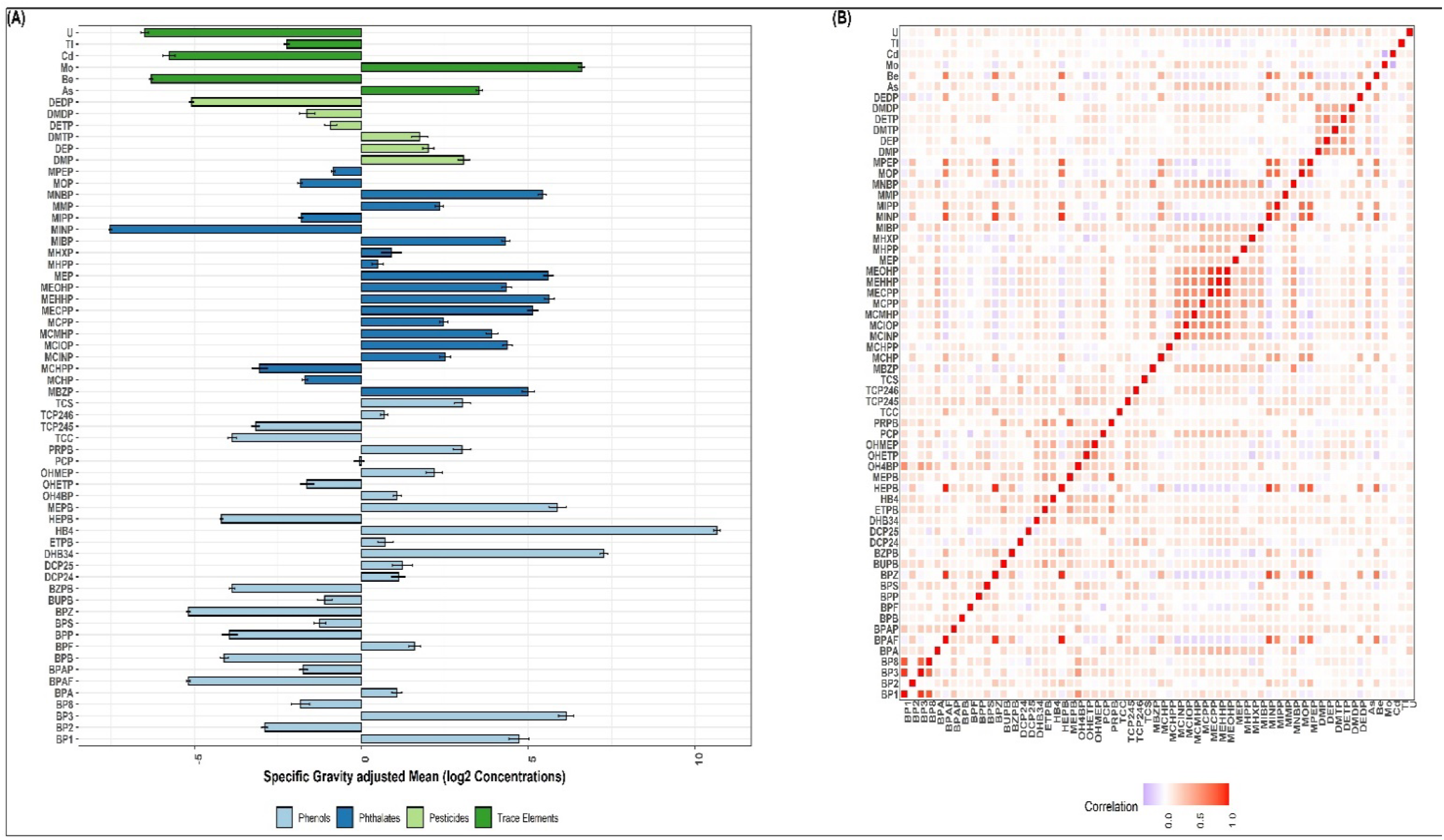
Specific gravity-adjusted and log-transformed (base 2) mean concentration and correlation plot of urinary chemicals

There were moderate to strong (0.3 to 0.7) within-group correlations among pesticides and phenols. The distributions of the child’s sex and race/ethnicity were not significantly different between ASD and TD children (Table 1). Further, there was no significant difference in parental homeowner status. However, children with ASD were more likely to be older at their age of assessment, and their mothers were more likely to have any hypertensive disorder or diabetes for any BMI category. The chemical concentrations of Methyl Paraben, Diethyl-phosphate, and Propylparaben (the top three chemicals based on weights from WQS) were significantly higher in children with ASD.

**Table 1:** Characteristics of mothers and children included in the analysis from the CHARGE cohort.

| n = 479 | All (mean(SD)<br>or n(%)) | TD | ASD | p-value |
| --- | --- | --- | --- | --- |
| Child sex |  |  |  | 0.99 |
| Female | 91 (19) | 47 | 44 |  |
| Male | 388 (81) | 201 | 187 |  |
| Child Race/Ethnicity |  |  |  | 0.25 |
| White (non-Hispanic) | 246 (51.36) | 135 (54.44) | 111 (48.05) |  |
| Non-White (non-Hispanic) | 102 (21.29) | 46 (18.55) | 56 (24.24) |  |
| Hispanic any race | 131 (27.35) | 67 (27.02) | 64 (27.71) |  |
| Child age at assessment (in years) | 3.94 (0.75) | 3.82 (0.75) | 4.05 (0.73) | <0.01 |
| Child Year of Birth (baseline 2000)<sup>#</sup> | 6.86 (3.08) | 6.48 (2.91) | 7.26 (3.21) | <0.01 |
| Parental homeowner status |  |  |  | 0.09 |
| No | 137 (28.60) | 62 (25) | 75 (32.47) |  |
| Yes | 342 (71.40) | 186 (75) | 156 (67.53) |  |
| Maternal Age child's birth | 30.57 (5.56) | 30.42 (5.43) | 30.73 (5.71) | 0.38 |
| Maternal Metabolic Condition<sup>##</sup> |  |  |  | 0.01 |
| Healthy (BMI <25) weight and no metabolic conditions | 230 (48.02) | 124 (50.00) | 106 (45.89) |  |
| verweight (BMI: 25-29.9) and no metabolic conditions | 102 (21.29) | 60 (24.19) | 42 (18.18) |  |
| Obese (BMI >30), no other metabolic conditions | 68 (14.20) | 36 (14.52) | 32 (13.85) |  |
| Any hypertensive disorder (including obesity) or diabetes | 79 (16.49) | 28 (11.29) | 51 (22.08) |  |
| MEPB** (in ng/ml) | 5.87 (2.87) | 5.47 (2.81) | 6.31 (2.87) | <0.01 |
| DEP** (in ng/ml) | 2.01 (1.78) | 1.76 (1.64) | 2.27 (1.89) | <0.01 |
| PRPB** (in ng/ml) | 3.02 (2.96) | 2.66 (2.94) | 3.39 (2.95) | <0.01 |

\*transformed to log (base 2) and corrected for specific gravity, \*\*top three chemicals in terms of weights from WQS regression, <sup>#</sup>all children were born after 2000, <sup>##</sup> the covariate maternal metabolic condition was created in previous studies by merging BMI categories with any hypertensive disorder and obesity and was shown to be associated with neuro-developmental outcomes in children<sup>69</sup>. P-values for the difference between ASD and TD groups were calculated using the Fisher exact test for categorical variables and the Wilcoxon rank-sum test for continuous variables. ASD: Autism Spectrum Disorder, TD: Typical Development, BMI: Body Mass Index, MEPB: Methyl Paraben, DEP: Diethyl-phosphate, PRPB: Propylparaben

### WQS- SiRF result

In the WQS model (with binary outcome ASD vs. TD and without any interaction term), the mixture index was significantly associated with higher odds of ASD (OR[95% CI]: 1.58[1.32, 1.88]). There were 20 chemicals with higher than chance contribution (weight > 1/62) to the overall mixture effect. The top five chemicals were Methyl Paraben, Diethyl-phosphate, Propylparaben, trace-metal Uranium, and Bisphenol F. The estimated weights (and the corresponding 95% CIs) were presented in Figure 3.

**Figure 3:**
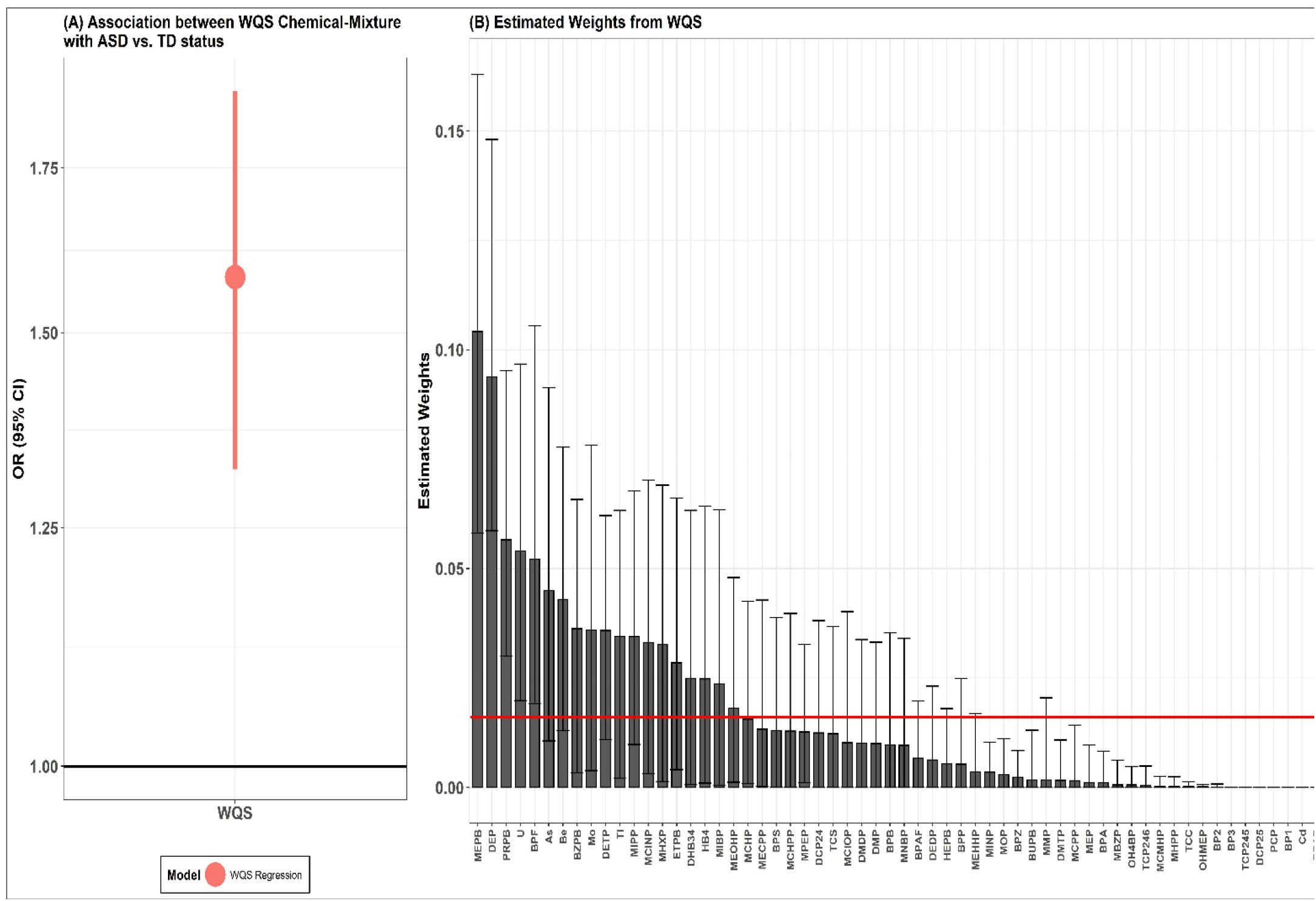
Results from WQS chemical mixture regression. (A) the overall OR between the WQS chemical mixture with ASD vs. TD status and (B) the corresponding weights contributed to the overall OR (note that the weights sum up to 1).

WQS-SiRF searched for interactions of multiple orders (>= 2) but eventually found two synergistic two-ordered interactions with more than 75% stability. The interactions were 1) urinary trace element Cadmium (Cd) and alkyl-phosphate pesticide – Diethyl-phosphate (DEP), denoted by Cd/DEP; and 2) environmental Phenol 2,4,6-Trichlorophenol (TCP-246) and DEP, denoted by TCP-246/DEP. However, both interactions were only observed in a subset of the sample whose urinary chemical concentrations of Cd, DEP, and TCP-246 were higher than certain thresholds. Therefore, based on a 75^th^ percentile threshold cutoff, we created two separate interaction indicators to test these discovered interactions for association analysis. For example, if both the specific gravity-adjusted concentrations of Cd and DEP were more than the 75^th^ percentile, then the interaction indicator Cd/DEP would be non-zero; else, it would be zero. In the sample, the estimated prevalence of these interactions was 5% and 8.4% for Cd/DEP and TCP-246/DEP, respectively. The results of SiRF from all three different data partitions were presented in Supplemental Table S4.

In two separate adjusted models (after controlling for the main WQS chemical mixture and covariates), each interaction indicator was associated with increased odds of ASD, 2.60[0.90, 7.50] and 1.14[0.55, 2.38] for Cd/DEP and TCP-246+/DEP respectively. Find all the ORs and corresponding CIs in the forest plot in Figure 4. Among the two interactions, Cd/DEP had the strongest association, and in all the models, the WQS chemical mixture remained statistically significant, with just a slight change in the ORs.

**Figure 4:**
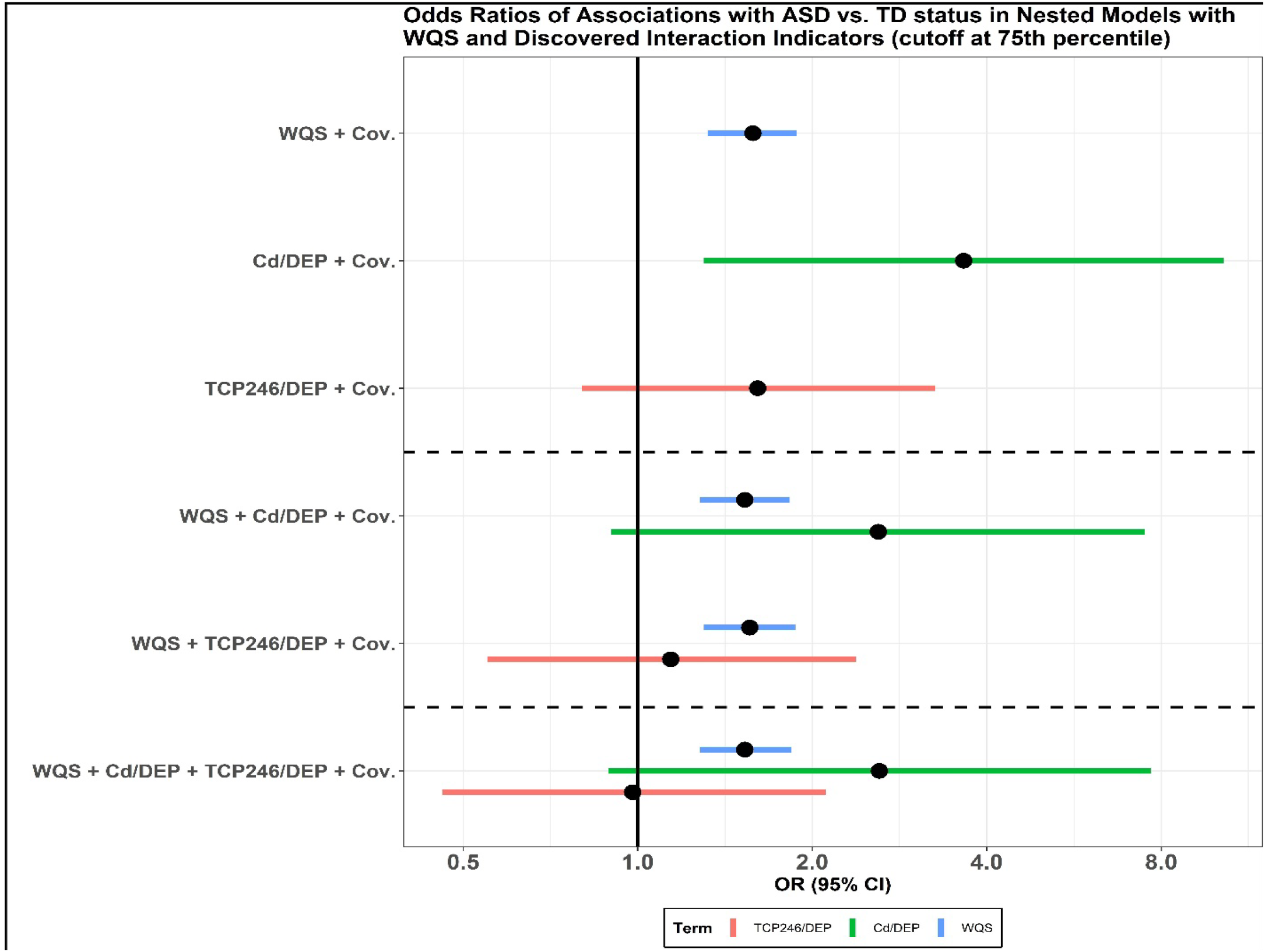
Results from nested linear models with WQS and discovered interaction indicators (cutoff set at 75^th^ percentile) and WQS chemical mixture. In the top three models, the WQS chemical mixture and the two interaction indicators were used in separate models. Both interaction indicators were adjusted with the WQS chemical mixture in the following two models. In the last model, interaction indicators and the WQS chemical mixture were put in the same model. All models were adjusted for covariates.

In the sensitivity analyses, (1) the interactions Cd/DEP and TCP-246/DEP were replicated when the WQS-SiRF algorithm was re-fitted without chemicals whose % of detection above LOD was less than 60% (Supplemental Table S5). (2) Furthermore, the gradual increase in the number of bootstraps, from 250, 500, to 1000, did not alter the results. Both the discovered interactions remained unaltered when the whole dataset was used to test the model trained on 75% data (Supplemental Table S6). Moreover, (3) the directionality of the ORs did not alter even when the interaction threshold of the 75^th^ percentile was changed to the 67^th^ percentile (Supplemental Figure S1), and (4) the interactions Cd/DEP and TCP-246/DEP were not found in the permutation tests.

## Discussion

We leveraged data from the CHARGE study to assess the synergistic interactions between environmental chemicals, pesticides, phthalates, phenols, and trace elements and ASD. Utilizing WQS-SiRF, we found two suggestive synergistic interactions associated with increased odds of ASD diagnosis between (1) Cd and DEP and (2) 2,4,6-trichlorophenol and DEP among children with the urinary concentration of interacting chemicals over certain thresholds. When controlled for the main WQS mixture and the necessary covariates, cadmium/DEP and TCP-246/DEP were associated with increased odds of ASD, respectively. Between the two interactions, cadmium/DEP had the strongest association and was previously shown to form chemical complexes. The identified interactions could be experimentally testable and, therefore, biologically meaningful. This paper is a continuation of the study of the main effects by Bennet et al.^13^, which concluded that many urinary chemicals were associated with increased odds of ASD at 2-5 years of age. The present study adds value by examining multi-ordered synergistic interactions between exposures to pesticides, phthalates, phenols, and trace elements and ASD and providing evidence for suggestive synergistic interactions observed between Cd/DEP and TCP-246/DEP.

There are few studies on interactions associated with ASD, including gene-environment^70^, social^71^, and chemical^4^ factors. Moreover, there is a lack of studies demonstrating chemical-chemical interactions in this context. Previous studies have shown an association between heavy metals, like cadmium, and ASD^72, 73^. Kern et al. discovered that cadmium and other trace elements were significantly lower in the hair of children with autism than others^74^. This supports the concept that children with autism may have issues excreting cadmium, resulting in a higher body burden that could contribute to symptoms of autism^74, 75^. Children could be exposed to cadmium through inhalation and ingestion. It is commonly found in the food chain, soil, cigarette smoke, and manufactured products^73^. Research on pesticide exposure during childhood, specifically glyphosate^76, 77^, chlorpyrifos^77^, diazinon^77^, and the development of ASD continues to emerge^78-80^. Potential routes of pesticide exposure in children include food contaminated with pesticides (ingestion), in utero or through breastmilk, and household exposures via dermal contact^81, 82^. However, there is a lack of studies showing any associations between the interaction of DEP and TCP-246 with ASD. Regarding possible biochemical significance, the cation, Cd^2+^ forms a complex with phosphate ester, particularly with DEP (C_4_H_10_O_4_P^-^), forming cadmium diethyl phosphate, C_4_H_10_CdO_4_P^-^^83, 84^. Although for the TCP-246+/DEP+ interaction, many details are not known, a chemical complex “2,4,6-trichlorophenyl dialkyl phosphate” was patented (in 1952) for use as parasiticides and control of agricultural and household pests through aqueous suspensions employed as sprays^85^. However, the activities of both chemical complexes in biological media are not known in detail.

We acknowledge the study’s limitations. (1) The urine samples were collected post-diagnosis, i.e., months and sometimes years after their symptoms emerged. In rare cases, urine samples were collected at the diagnosis. Therefore we cannot rule out reverse causation, with the associations reflecting lifestyle patterns. In addition, urinary measurements of the organic compounds assessed in this study represent recent exposures due to their half-lives. Ideally, the urine samples should have been assessed with repeated samples collected at various time points^13, 86-88^. As a result, we are uncertain whether these chemical interactions directly contribute to ASD diagnosis. (2) Because of the limited sample size, we did not study potential sex-specific associations with ASD diagnosis, although sexually dimorphic effects are well documented.^2^ (3) Additionally, we used the same confounders used in the original analysis by Bennet et al.^13^. However, these confounders were selected based on them being confounders to MEPB because it has one of the strongest associations in the unadjusted model. (4) Similar to large case-control studies, residual confounding is possible. However, our results remained unaltered after adjusting for multiple confounders and covariates, negating residual confounding as the sole explanation. (5) The choice of cutoffs at the 75^th^ or 67^th^ percentile is ad-hoc and sample-specific and therefore needs to be replicated in a separate independent study population. Further, using random intersection trees within the SiRF algorithm makes it difficult to extract the absolute threshold cutoffs directly. Therefore given these methodological challenges, there lies a strong potential for further developments attuned to specific problems. (6) In the present analysis, the same chemicals were used in the WQS and then again in the SiRF, raising the possibility of over-fitting. A training, testing, and validation data split in an ideal large sample scenario would potentially guard against overfitting. However, in this moderate sample-sized study, the use of random subsets and repeated holdouts in training and testing samples of WQS and the drawing of a large number of bootstrapped samples with different training and testing splits in the SiRF could potentially induce a robust guard against overfitting.

However, our study has several strengths. First, CHARGE is a well-established case-control study with extensive demographic and covariate data. Further, it allowed us to assess a wide range of environmental chemical exposures in children 2-5 years of age, along with available data on ASD with a moderate sample size. Second, this is the first study to combine exposure mixture methods and machine learning tools to discover interactions that mimic classical threshold-based toxicological dose-response interactions, providing a meaningful way to extract plausible mechanistic insight. Third, these toxicologically mimicking interactions are only present in a subset of the sample, therefore, can be thought of as “personalized and precision” interactions. Fourth, WQS-SiRF can efficiently search for high-order interactions; therefore, the intended order does not need to be specified beforehand. Fifth, in terms of practical implementation, the WQS-SiRF algorithm is relatively fast and user-friendly, with both having robust R packages. In conclusion, we introduced a novel way of discovering threshold-based interactions. To the best of our knowledge, this is the first paper that combines the inferential power of WQS and the predictive accuracy of a machine-learning algorithm to discover threshold-based personalized biologically suggestive interactions among urinary biomarkers associated with higher odds of ASD.

## Data Availability

All data produced are available online at Human Health Exposure Analysis Resource (HHEAR) Data Center at the Icahn School of Medicine at Mount Sinai

https://hheardatacenter.mssm.edu/

## Acknowledgment

We want to thank the Human Health Exposure Analysis Resource (HHEAR) Data Center at the Icahn School of Medicine at Mount Sinai for the availability of open-source data and the CHARGE study participants and researchers for making this work possible.

