## Supplementary Information for "Machine learning assisted discovery of synergistic interactions between environmental pesticides, phthalates, phenols, and trace elements in child neurodevelopment"

**Figure S1**

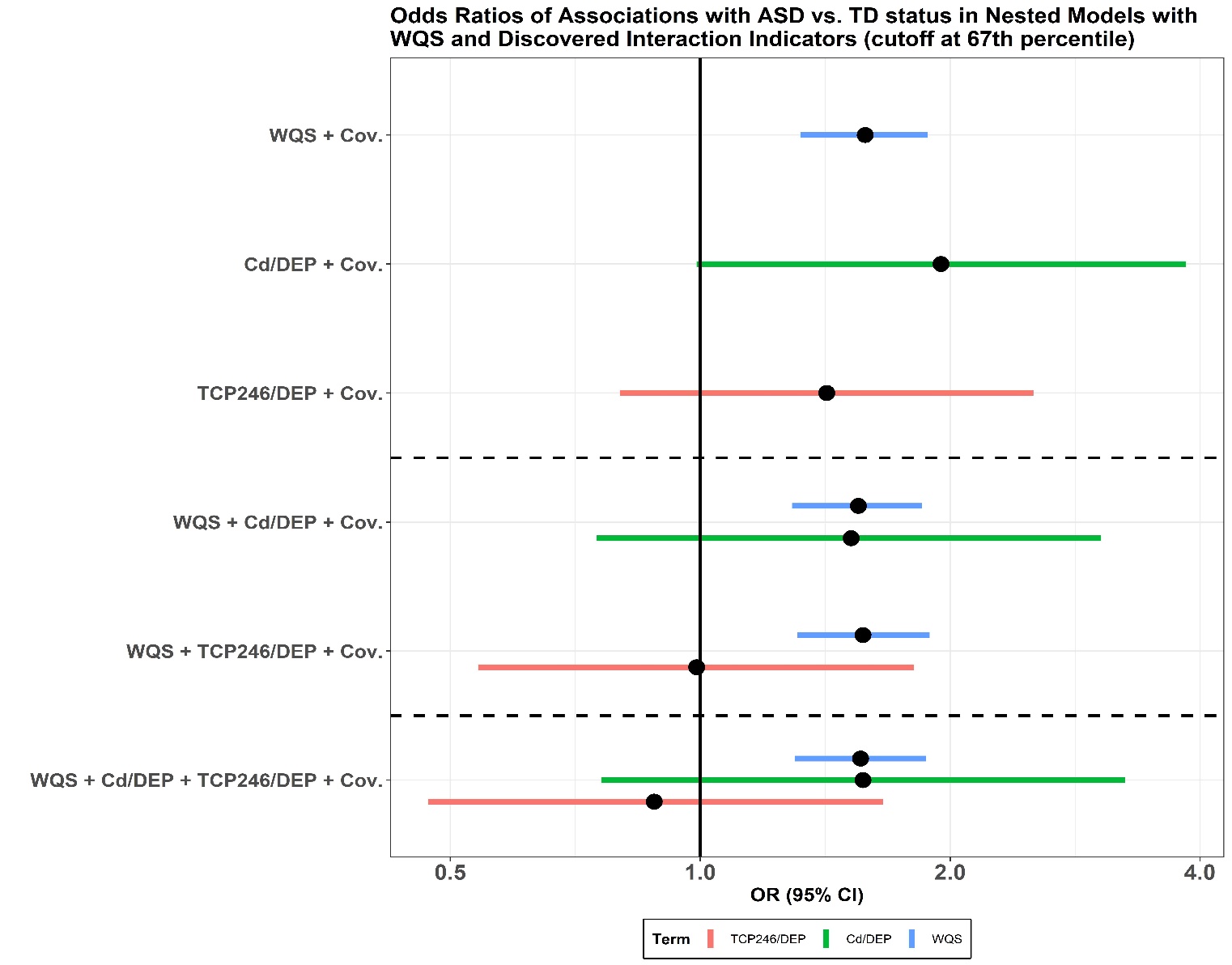

**Figure S1:** Results from nested linear models with WQS and discovered interaction indicators (cutoff set at 67^th^ percentile) and WQS chemical mixture. In the top three models, the WQS chemical mixture and the two interaction indicators were used in separate models. Both interaction indicators were adjusted for the WQS chemical mixture in the following two models. In the last model, interaction indicators and the WQS chemical mixture were put in the same model. All models were adjusted for covariates.

**Table S1: Chemical names and abbreviations***

| Chemical class | Chemical Abbreviation | Chemical Full Name |
| --- | --- | --- |
| Phenols and Parabens | BP1 | Benzophenone-1 |
|  | BP2 | 2,2',4,4'-tetrahydroxybenzophenone |
|  | BP3 | Benzophenone-3 |
|  | BP8 | 2,2'-dihydroxy-methoxybenzophenone |
|  | BPA | Bisphenol A |
|  | BPAF | 4,4'-(hexafluoroisopropylidene)-dephenol |
|  | BPAP | 4,4'-(1-phenylethylidene)bisphenol |
|  | BPB | 2,2'-bis(4-hydroxyphenyl)butane |
|  | BPF | Bisphenol F |
|  | BPP | 4,4′-(1,4-Phenylenediisopropylidene)bisphenol |
|  | BPS | Bisphenol S |
|  | BPZ | 4,4'-cyclo-hexylidenebisphenol |
|  | BUPB | Butyl paraben |
|  | BZPB | Benzyl paraben |
|  | DCP24 | 2,4-Dichlorophenol |
|  | DCP25 | 2,5-Dichlorophenol |
|  | DHB34 | 3,4-Dihydroxy benzoic acid |
|  | ETPB | Ethyl paraben |
|  | HB4 | 4-Hydroxybenzoic acid |
|  | HEPB | Heptyl paraben |
|  | MEPB | Methyl Paraben |
|  | OH4BP | 4-hydroxybenzophenone |
|  | OHETP | Protocatechuic acid ethyl ester |
|  | OHMEP | 3,4-Dihydroxy-benzoic acid methyl ester |
|  | PCP | Pentachlorophenol |
|  | PRPB | Propyl paraben |
|  | TCC | Triclocarban |
|  | TCP245 | 2,4,5-Trichlorophenol |
|  | TCP246 | 2,4,6-Trichlorophenol |
|  | TCS | Triclosan |
| Phthalates | MBZP | mono-benzyl phthalate |
|  | MCHP | Monocyclohexyl phthalate |
|  | MCHPP | mono(7-carboxyheptyl)phthalate |
|  | MCINP | mono-carboxy isononyl phthalate |
|  | MCIOP | mono-carboxy isooctyl phthalate |
|  | MCMHP | Mono-2-(carboxymethyl) hexyl phthalate |
|  | MCPP | mono (3-carboxypropyl) phthalate (multiple) |
|  | MECPP | mono-(2-ethyl-5-carboxypentyl) phthalate |
|  | MEHHP | mono (2-ethyl-5-hydroxyhexyl) phthalate |
|  | MEOHP | mono (2-ethyl-5-oxohexyl) phthalate |
|  | MEP | monoethyl phthalate (DEP) |
|  | MHPP | mono-2-heptyl phthalate |
|  | MHXP | mono-hexyl phthalate |
|  | MIBP | mono-isobutyl phthalate (DiBP) |
|  | MINP | mono-isononyl phthalate |
|  | MIPP | Mono-isopropyl phthalate |
|  | MMP | mono-methyl phthalate |
|  | MNBP | mono-n-butylphthalate (BzBP, DnBP) |
|  | MOP | Mono-n-octyl phthalate |
|  | MPEP | mono-pentyl phthalate |
| Pesticides | DMP | Dimethylphosphate |
|  | DEP | Diethylphosphate |
|  | DMTP | Dimethylthiophosphate |
|  | DETP | Diethylthiophosphate |
|  | DMDP | Dimethyldithiophosphate |
|  | DEDP | Diethyldithiophosphate |
| Trace Elements | As | Arsenic |
|  | Be | Beryllium |
|  | Cd | Cadmium |
|  | Mo | Molybdenum |
|  | Tl | Thallium |
|  | U | Uranium |

* From (Bennett et al. 2022)

**Table S2: Limit of detection (LOD) for individual chemicals and percent detection above the LOD by chemical classes**

| Phenols (N=479) | | | Phthalates (N=479) | | | Pesticides (N=479) | | | Trace Elements (N=479) | | |
| --- | --- | --- | --- | --- | --- | --- | --- | --- | --- | --- | --- |
| Chemical | LOD | % above LOD | Chemical | LOD (ng/ml) | % above LOD | Chemical | LOD (ng/ml) | % above LOD | Chemical | LOD (ng/ml) | % above LOD |
| BP1 | 0.05 | 100 | MBZP | 0.02 | 99.25 | DMP | 0.05 | 99.55 | As | 0.059 | 100 |
| BP2 | 0.2 | 11.19 | MCHP | 0.50 | 07.46 | DEP | 0.10 | 99.25 | Be | 0.022 | 6.57 |
| BP3 | 0.5 | 100 | MCHPP | 0.10 | 26.57 | DMTP | 0.05 | 95.82 | Mo | 0.45 | 100 |
| BP8 | 0.02 | 80.90 | MCINP | 0.01 | 98.66 | DETP | 0.02 | 98.51 | Cd | 0.006 | 72.84 |
| BPA | 0.05 | 98.66 | MCIOP | 0.01 | 99.85 | DMDP | 0.02 | 91.79 | Tl | 0.005 | 100 |
| BPAF | 0.05 | 0.75 | MCMHP | 0.02 | 98.81 | DEDP | 0.05 | 3.73 | U | 0.0007 | 99.55 |
| BPAP | 0.1 | 89.25 | MCPP | 0.05 | 99.85 |  |  |  |  |  |  |
| BPB | 0.05 | 50.75 | MECPP | 0.02 | 99.70 |  |  |  |  |  |  |
| BPF | 0.5 | 89.40 | MEHHP | 0.20 | 100 |  |  |  |  |  |  |
| BPP | 0.01 | 76.72 | MEOHP | 0.01 | 100 |  |  |  |  |  |  |
| BPS | 0.05 | 94.78 | MEP | 0.10 | 100 |  |  |  |  |  |  |
| BPZ | 0.05 | 1.19 | MHPP | 0.50 | 74.18 |  |  |  |  |  |  |
| BUPB | 0.05 | 92.84 | MHXP | 0.50 | 57.16 |  |  |  |  |  |  |
| BZPB | 0.1 | 14.48 | MIBP | 0.01 | 100 |  |  |  |  |  |  |
| DCP24 | 0.02 | 95.07 | MINP | 0.01 | 0 |  |  |  |  |  |  |
| DCP25 | 0.02 | 88.06 | MIPP | 0.50 | 2.69 |  |  |  |  |  |  |
| DHB34 | 0.2 | 100 | MMP | 5.0 | 41.34 |  |  |  |  |  |  |
| ETPB | 0.02 | 99.1 | MNBP | 0.20 | 99.85 |  |  |  |  |  |  |
| HB4 | 1 | 100 | MOP | 0.50 | 1.64 |  |  |  |  |  |  |
| HEPB | 0.1 | 0.30 | MPEP | 1.0 | 2.69 |  |  |  |  |  |  |
| MEPB | 0.05 | 99.85 |  |  |  |  |  |  |  |  |  |
| OH4BP | 0.2 | 98.81 |  |  |  |  |  |  |  |  |  |
| OHETP | 0.1 | 65.67 |  |  |  |  |  |  |  |  |  |
| OHMEP | 0.1 | 96.12 |  |  |  |  |  |  |  |  |  |
| PCP | 0.05 | 96.27 |  |  |  |  |  |  |  |  |  |
| PRPB | 0.1 | 98.51 |  |  |  |  |  |  |  |  |  |
| TCC | 0.1 | 07.91 |  |  |  |  |  |  |  |  |  |
| TCP245 | 0.1 | 51.75 |  |  |  |  |  |  |  |  |  |
| TCP246 | 0.02 | 99.85 |  |  |  |  |  |  |  |  |  |
| TCS | 0.1 | 96.87 |  |  |  |  |  |  |  |  |  |

**Table S3: Distribution of specific gravity-adjusted urinary phenol, phthalate, and trace element biomarker concentrations (ng/ml) among 479 participants included in the analysis**

| **Chemical** | **5^th^ Percentile** | **25^th^ Percentile** | **Median** | **75^th^ Percentile** | **95^th^ Percentile** |
| --- | --- | --- | --- | --- | --- |
| ***Phenols*** |  | | | | |
| **BP1** | 0.76 | 2.58 | 4.00 | 5.91 | 12.75 |
| **BP2** | -3.84 | -3.53 | -3.16 | -2.54 | -0.96 |
| **BP3** | 2.75 | 4.32 | 5.82 | 7.64 | 10.51 |
| **BP8** | -6.75 | -3.51 | -1.87 | -0.10 | 3.45 |
| **BPA** | -1.53 | 0.15 | 1.16 | 1.98 | 3.52 |
| **BPAF** | -5.87 | -5.60 | -5.30 | -4.82 | -4.32 |
| **BPAP** | -4.09 | -2.58 | -1.64 | -0.88 | 0.40 |
| **BPB** | -5.70 | -5.16 | -4.32 | -3.24 | -1.94 |
| **BPF** | -2.22 | 0.41 | 1.65 | 3.07 | 4.44 |
| **BPP** | -7.92 | -6.39 | -3.50 | -2.14 | -0.38 |
| **BPS** | -4.32 | -2.56 | -1.30 | -0.09 | 2.03 |
| **BPZ** | -5.87 | -5.60 | -5.30 | -4.82 | -4.32 |
| **BUPB** | -4.46 | -2.51 | -1.34 | 0.03 | 3.75 |
| **BZPB** | -4.84 | -4.54 | -4.13 | -3.43 | -1.90 |
| **DCP24** | -4.38 | 0.32 | 1.40 | 2.48 | 3.68 |
| **DCP25** | -6.66 | 0.61 | 1.83 | 2.73 | 5.88 |
| **DHB34** | 5.17 | 6.48 | 7.39 | 8.10 | 9.12 |
| **ETPB** | -2.13 | -0.84 | 0.32 | 1.68 | 5.62 |
| **HB4** | 9.20 | 10.06 | 10.55 | 11.24 | 12.75 |
| **HEPB** | -4.87 | -4.60 | -4.30 | -3.84 | -3.32 |
| **MEPB** | 2.14 | 3.63 | 5.41 | 7.55 | 11.29 |
| **OH4BP** | -1.11 | 0.35 | 0.97 | 1.67 | 3.71 |
| **OHETP** | -4.62 | -3.57 | -1.51 | -0.02 | 1.97 |
| **OHMEP** | -2.28 | 0.63 | 2.16 | 3.71 | 6.61 |
| **PCP** | -2.53 | -0.75 | 0.05 | 0.91 | 2.14 |
| **PRPB** | -1.16 | 0.95 | 2.60 | 4.73 | 8.52 |
| **TCC** | -4.85 | -4.58 | -4.20 | -3.65 | -1.51 |
| **TCP245** | -4.70 | -4.16 | -3.32 | -2.38 | -0.89 |
| **TCP246** | -1.07 | -0.01 | 0.56 | 1.28 | 2.61 |
| **TCS** | -1.34 | 1.49 | 2.98 | 4.72 | 7.42 |

| **Chemical** | **5^th^ Percentile** | **25^th^ Percentile** | **Median** | **75^th^ Percentile** | **95^th^ Percentile** |
| --- | --- | --- | --- | --- | --- |
| ***Phthalates*** |  | | | | |
| **MBZP** | 2.15 | 3.88 | 4.97 | 6.13 | 8.04 |
| **MCHP** | -2.52 | -2.25 | -1.88 | -1.33 | -0.34 |
| **MCHPP** | -4.84 | -4.49 | -3.93 | -3.29 | 3.90 |
| **MCINP** | 0.47 | 1.81 | 2.54 | 3.30 | 4.84 |
| **MCIOP** | 2.07 | 3.59 | 4.37 | 5.20 | 6.81 |
| **MCMHP** | 1.20 | 3.13 | 4.08 | 5.08 | 6.51 |
| **MCPP** | 0.40 | 1.66 | 2.45 | 3.27 | 4.50 |
| **MECPP** | 2.97 | 4.27 | 5.04 | 6.06 | 7.59 |
| **MEHHP** | 3.32 | 4.64 | 5.48 | 6.50 | 8.34 |
| **MEOHP** | 1.98 | 3.30 | 4.24 | 5.33 | 7.03 |
| **MEP** | 3.64 | 4.62 | 5.41 | 6.31 | 8.34 |
| **MHPP** | -2.11 | -1.00 | 0.43 | 1.69 | 3.72 |
| **MHXP** | -2.38 | -1.73 | -0.56 | 3.56 | 7.12 |
| **MIBP** | 2.32 | 3.47 | 4.31 | 5.14 | 6.59 |
| **MINP** | -8.19 | -7.92 | -7.62 | -7.16 | -6.64 |
| **MIPP** | -2.52 | -2.28 | -1.95 | -1.43 | -1.00 |
| **MMP** | 0.89 | 1.34 | 2.10 | 2.98 | 4.84 |
| **MNBP** | 3.53 | 4.65 | 5.45 | 6.19 | 7.39 |
| **MOP** | -2.55 | -2.28 | -1.95 | -1.48 | -1.00 |
| **MPEP** | -1.52 | -1.28 | -0.95 | -0.48 | 0.00 |

| **Chemical** | **5^th^ Percentile** | **25^th^ Percentile** | **Median** | **75^th^ Percentile** | **95^th^ Percentile** |
| --- | --- | --- | --- | --- | --- |
| ***Pesticides*** |  | | | | |
| **DMP** | 0.22 | 1.82 | 3.16 | 4.34 | 6.06 |
| **DEP** | -0.63 | 0.88 | 2.02 | 3.18 | 4.78 |
| **DMTP** | -3.39 | 0.19 | 1.81 | 3.68 | 5.62 |
| **DETP** | -4.10 | -2.25 | -1.03 | 0.32 | 2.52 |
| **DMDP** | -6.12 | -3.23 | -1.69 | 0.01 | 2.99 |
| **DEDP** | -5.85 | -5.57 | -5.24 | -4.75 | -4.32 |

| **Chemical** | **5^th^ Percentile** | **25^th^ Percentile** | **Median** | **75^th^ Percentile** | **95^th^ Percentile** |
| --- | --- | --- | --- | --- | --- |
| ***Trace Elements*** |  | | | | |
| **As** | 1.99 | 2.83 | 3.47 | 4.11 | 5.57 |
| **Be** | -7.03 | -6.77 | -6.42 | -5.86 | -5.51 |
| **Mo** | 5.02 | 6.04 | 6.62 | 7.18 | 8.25 |
| **Cd** | -8.81 | -7.43 | -5.46 | -4.37 | -3.03 |
| **Tl** | -3.53 | -2.71 | -2.25 | -1.78 | -0.98 |
| **U** | -8.48 | -7.35 | -6.62 | -5.68 | -4.26 |

**Table S4: Results of SiRF from the three different data partitions**

- **70% training and 30% testing data**

| **Interactions** | **Prevalence** | **Precision** | **Stability** |
| --- | --- | --- | --- |
| DEP+_U+ | 0.069 | 0.555 | 0.884 |
| DEP+_TCP246+ | 0.065 | 0.473 | 0.568 |
| Cd+_DEP+ | 0.109 | 0.447 | 0.944 |
| BPAP+_DEP+ | 0.115 | 0.448 | 0.928 |

- **75% training and 25% testing data**

| **Interactions** | **Prevalence** | **Precision** | **Stability** |
| --- | --- | --- | --- |
| DEP+_Mo+ | 0.076 | 0.558 | 0.936 |
| DEP+_MEPB+ | 0.053 | 0.573 | 0.696 |
| DEP+_U+ | 0.049 | 0.547 | 0.620 |
| DEP+_TCP246+ | 0.083 | 0.480 | 0.792 |
| Cd+_DEP+ | 0.100 | 0.460 | 0.848 |
| BPAP+_DEP+ | 0.117 | 0.449 | 0.928 |

- **80% training and 20% testing data**

| **Interactions** | **Prevalence** | **Precision** | **Stability** |
| --- | --- | --- | --- |
| DEP+_MEPB+ | 0.052 | 0.565 | 0.672 |
| DEP+_TCP246+ | 0.098 | 0.484 | 0.908 |
| Cd+_DEP+ | 0.117 | 0.464 | 0.956 |
| Cd+_TCP246+ | 0.078 | 0.448 | 0.512 |

**Table S5: Results of SiRF from the three different data partitions without chemicals whose % of detection above LOD was less than 60%**

- **70% training and 30% testing data**

| **Interactions** | **Prevalence** | **Precision** | **Stability** |
| --- | --- | --- | --- |
| DEP+_U+ | 0.062 | 0.553 | 0.868 |
| DEP+_DETP+ | 0.055 | 0.506 | 0.660 |
| DEP+_TCP246+ | 0.067 | 0.474 | 0.624 |
| DEP+_MCMHP+ | 0.074 | 0.447 | 0.708 |
| Cd+_DEP+ | 0.085 | 0.445 | 0.756 |
| BPAP+_DEP+ | 0.108 | 0.443 | 0.884 |

- **75% training and 25% testing data**

| **Interactions** | **Prevalence** | **Precision** | **Stability** |
| --- | --- | --- | --- |
| DEP+_Mo+ | 0.069 | 0.564 | 0.864 |
| DEP+_MEPB+ | 0.049 | 0.566 | 0.664 |
| DEP+_U+ | 0.042 | 0.542 | 0.528 |
| DEP+_Tl+ | 0.051 | 0.527 | 0.576 |
| DEP+_TCP246+ | 0.078 | 0.480 | 0.764 |
| DEP+_MCPP+ | 0.052 | 0.461 | 0.504 |
| DEP+_MCMHP+ | 0.061 | 0.449 | 0.596 |
| Cd+_DEP+ | 0.078 | 0.456 | 0.708 |
| BPAP+_DEP+ | 0.094 | 0.444 | 0.792 |

- **80% training and 20% testing data**

| **Interactions** | **Prevalence** | **Precision** | **Stability** |
| --- | --- | --- | --- |
| DEP+_TCP246+ | 0.086 | 0.485 | 0.808 |
| Cd+_DEP+ | 0.093 | 0.461 | 0.856 |
| DEP+_MCMHP+ | 0.071 | 0.444 | 0.628 |
| BPAP+_DEP+ | 0.090 | 0.450 | 0.724 |

**Table S6: Results of SiRF when the whole dataset was used to test the model trained on 75% data**

| **Interactions** | **Prevalence** | **Precision** | **Stability** |
| --- | --- | --- | --- |
| DEP+_Mo+ | 0.076 | 0.558 | 0.936 |
| DEP+_MEPB+ | 0.053 | 0.573 | 0.696 |
| DEP+_U+ | 0.049 | 0.547 | 0.620 |
| DEP+_TCP246+ | 0.083 | 0.480 | 0.792 |
| Cd+_DEP+ | 0.100 | 0.460 | 0.848 |
| BPAP+_DEP+ | 0.117 | 0.449 | 0.928 |

**Tuning Parameters for WQS-SiRF**

1. **WQS:** q = 10, signal = "t2", b = 500, rs = T, n_vars = 12, rh = 100, validation = 0.25, seed = 123123123, b1_pos = T, family = "binomial", plan_strategy = "multicore"
2. **SiRF:** seed was set at set.seed(123456) and n.iter=5, n.core=1, select.iter = T, n.bootstrap=500
3. **Selection of training and testing data**

- **70% training and 30% testing data:**

set.seed(123456)

train.id <- sample(seq(1,n), ceiling(n*70/100))

test.id <- setdiff(1:n, train.id)

- **75% training and 25% testing data:**

set.seed(123456)

train.id <- sample(seq(1,n), ceiling(n*75/100))

test.id <- setdiff(1:n, train.id)

- **80% training and 20% testing data:**

set.seed(123456)

train.id <- sample(seq(1,n), ceiling(n*80/100))

test.id <- setdiff(1:n, train.id)
